# Global transmission suitability maps for dengue virus transmitted by *Aedes aegypti* from 1981 to 2019

**DOI:** 10.1101/2022.11.04.22281958

**Authors:** Taishi Nakase, Marta Giovanetti, Uri Obolski, José Lourenço

## Abstract

Mosquito-borne viruses increasingly threaten human populations due to accelerating changes in climate, human and mosquito migration, and land use practices. Over the last three decades, the global distribution of dengue has rapidly expanded, causing detrimental health and economic problems in many areas of the world. To develop effective disease control measures and plan for future epidemics, there is an urgent need to map the current and future transmission potential of dengue across both endemic and emerging areas. Expanding and applying Index P, a previously developed mosquito-borne viral suitability measure, we map the global climate-driven transmission potential of dengue virus transmitted by *Aedes aegypti* mosquitoes from 1981 to 2019. This database of dengue transmission suitability maps and an R package for Index P estimations are offered to the public health community as resources towards the identification of past, current and future transmission hotspots. These resources and the studies they facilitate can contribute to the planning of disease control and prevention strategies, especially in areas where surveillance is unreliable or non-existent.

## Background & Summary

Global changes in environmental conditions that favor a closer proximity between vector and human populations are facilitating the spread of mosquito-borne viruses (MBVs)^1^. These processes, which include climate change, urbanization, deforestation and migration are interacting and evolving, creating a complex landscape of current and future disease risk. For example, the recent emergence and epidemic spread of Zika virus (ZIKV) caused severe health and economic problems in Latin America^2^. The public health impact of chikungunya (CHIKV) has also significantly increased in recent years with its rapid emergence in the Americas in late 2013^3–5^. The threat of these events is further magnified by the potential of their pathogens to cause severe health complications such as dengue haemorrhagic fever, ZIKV-induced congenital and neurological disorders, and CHIKV-associated Guillain-Barré Syndrome^6, 7^.

Dengue virus (DENV), transmitted by *Aedes spp*. mosquitoes, accounts for an increasingly large proportion of all vector-borne disease, with an estimated 400 million infections per year^8, 9^. The geographical range of DENV has expanded in recent years, becoming endemic in much of Central America, South America and Southeast Asia and establishing epidemic cycles in parts of Africa and North America^10^. In the last decade, Europe has also seen a rise in autochthonous DENV transmission as mosquito populations advance north from the Mediterranean and travel to endemic areas increases^11^. The only licensed dengue vaccine remains in limited use because of the higher risk of severe disease from breakthrough infections in seronegative recipients^12^. Current control efforts are focused on mosquito control and viral surveillance in human and mosquito populations^13^. Given the increasing threat posed by DENV and the reliance on vector control to mitigate transmission, understanding and mapping the transmission potential of DENV by *Aedes aegypti* mosquitoes, its primary vector, is essential for control and health systems planning.

The spatiotemporal dynamics of MBVs are primarily governed by the interplay among three factors: the physiological interactions between virus and vector, that between virus and host, and the population dynamics of the vector and host. The carrying capacity and seasonal oscillations of mosquito populations are influenced by many environmental and ecological factors including temperature^14–16^, humidity^16^, host population density^16, 17^, vegetation types^18^ and the presence of water sources^16, 17^. Climatic factors are particularly important because they also alter each mosquito’s potential to transmit the virus to new hosts by causing changes in physical and behavioral traits such as life span^19^, incubation period^20^ and biting rate^21^. With data from experimental studies, it is possible to mathematically characterize these relationships between climatic variables and mosquito-viral traits and incorporate them into mechanistic transmission models^22^.

The basic reproductive number (*R*_0_), which measures the average number of secondary infections generated by a single infectious host in a fully susceptible host population, is an important measure of transmission potential. For MBVs, expressions of *R*_0_ involve many interacting host, pathogen and vector variables, some of which are difficult to parameterize due to limited data (e.g. geographical mosquito distribution and density). To overcome this challenge, different measures of transmission potential, called suitability measures, have been developed^18, 21, 23, 24^. Vectorial capacity, one of the most widely known suitability measures, uses purely entomological variables to estimate the potential number of infectious bites that would arise from a single infectious person over one day^23^. A number of studies have built upon the concepts in vectorial capacity to develop new measures of transmission potential that consider the interactions between climate and mosquito-viral traits^22, 24, 25^. Here, we update and apply a previously developed mosquito-borne viral suitability measure^24^, referred to as Index P, to estimate geographical maps and time series for climate-driven DENV transmission potential of the *Ae. aegypti* vector. Index P is a proxy for the transmission potential of a single adult female mosquito under conditions where susceptible hosts, the virus and its vectors are assumed to be present^26^. Unlike previously developed suitability measures^18, 25^, Index P has a direct biological interpretation and takes into account factors that are not purely entomological (e.g. human infectious period 1*/σ*^*h*^ and the transmission probability from infected human to mosquito *ϕ*^*h*→*v*^). Local temperature and humidity time series are its main inputs, making its framework sufficiently general to be applied to any location for which such climate data exists and for any MBV for which there is empirical data on the relationship between climate and vector-viral traits. It has been successfully used to characterize the transmission potential and epidemiology of West Nile virus in Israel^27^, Portugal^28^, Brazil^29^, and Italy^30^, CHIKV and ZIKV in the Dominican Republic^5^ and Mexico^31^, and DENV in Myanmar^26^, Brazil^24, 32^and Mexico^31^. So far, the application of Index P has been performed at a small scale driven by specific and limited research goals. Given the sparsity of detailed epidemiological and ecological data for DENV (e.g. infection incidence and mosquito density) and the disproportionate impact of DENV in developing countries, there is a need to extend these analyses globally. Hence, we offer a database of spatiotemporal maps of Index P for 186 different countries and territories and an easy-to-use R package for Index P estimation as ready-to-use resources for the visualization and analysis of climate-driven DENV transmission potential globally over the last four decades.

In combination with previous work to map the distribution of DENV vectors^18, 33, 34^, these maps improve our understanding of past and current DENV transmission potential and have the capacity to inform disease control and prevention strategies. For example, in Central Africa where *Ae. aegypti* is predicted to be widely disseminated^18^ but reported DENV cases remain low^35^, these maps can help determine whether the apparent mismatch is due to low transmission potential of the vector or under-reporting and poor surveillance. This information can then be used to identify high-risk, low-surveillance areas where seroprevalence surveys should be targeted. It thus addresses a central limitation of occurrence-based predictions of dengue infection risk^8^, which can underestimate risk in areas where reported incidence is low or absent due to limited surveillance rather than low transmission. The maps may also be used to identify regions where *Ae. aegypti* might not be present but due to high DENV transmission potential there is a high likelihood of spillover of sylvatic DENV into human populations^36, 37^.

## Methods

### Fundamental theory of Index P

Index P is a climate-driven suitability measure for mosquito-borne viruses derived from a mechanistic model previously developed to study the transmission dynamics of Zika^38^ and dengue^26^. The equation for *R*_0_ in this model (equation (1)) can be decomposed into two components: the number of female mosquitoes per host (*M*) and the transmission potential of each female mosquito (*P*). Index P measures transmission potential solely based on *P* (equation (2)), which is interpreted as the reproductive potential of a single adult female mosquito in a fully susceptible host population. A description of the parameters in the equation for Index P can be found in Table 1. Given the availability of empirical studies that quantify the relationships between meteorological variables and viral-vector traits (e.g. extrinsic incubation period), it is possible to define equations for several parameters in the expression for *P* in terms of temperature (*t*) and humidity (*u*). Temperature and humidity time series data can be input into these equations to derive time series of the entomological parameters, which when combined with prior information on the selected vector/host/virus system can provide estimates of Index P over time.

**Table 1.** Descriptions and probability distributions of the biological parameters used in the estimation of Index P for DENV transmitted by Aedes aegypti mosquitoes. SD is standard deviation. The transmission probability per mosquito bite (*Ae. aegypti*-to-human and human-to-*Ae. aegypti*) are defined by deterministic temperature-dependent equations.

| Parameter | Symbol | Distribution | Mean, SD | Units | Sources |
| --- | --- | --- | --- | --- | --- |
| Adult *Ae. aegypti* lifespan | $1/\mu_{(u,t)}^v$ | normal | 10, 2.55 | days | 14, 19, 55 |
| Adult *Ae. aegypti* biting rate | $a_{(u)}^v$ | normal | 0.25, 0.01 | bites · mosq.<sup>-1</sup> · day<sup>-1</sup> | 56, 57 |
| Human lifespan | $1/\mu^h$ | normal | 70, 3 | years | - |
| Intrinsic human-DENV incubation period | $1/\gamma^h$ | lognormal | 5.94, 1.80 | days | 20 |
| Human-DENV infectious period | $1/\sigma^h$ | normal | 4, 0.51 | days | 58–60 |
| Extrinsic *Ae. aegypti*-DENV incubation period | $1/\gamma_{(t)}^v$ | lognormal | 1228, 1825 (∞, 15°C)<br>232, 181 [15°C, 17.5°C)<br>72.4, 32.6 [17.5°C, 20°C)<br>28.9, 7.64 [20°C, 22.5°C)<br>14.7, 2.66 [22.5°C, 25°C)<br>8.68, 1.31 [25°C, 27.5°C)<br>5.76, 0.87 [27.5°C, 30°C)<br>4.14, 0.63 [30°C, 32.5°C)<br>3.19, 0.48 [32.5°C, ∞) | days | 20 |
| Transmission probability per mosquito bite (*Ae. aegypti*-to-human) | $\phi_{(t)}^{v \rightarrow h}$ | none | N/A | N/A | N/A |
| Transmission probability per mosquito bite (human-to-*Ae. aegypti*) | $\phi_{(t)}^{h \rightarrow v}$ | none | N/A | N/A | N/A |

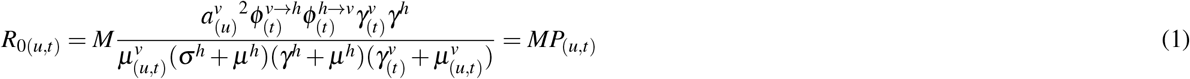

where

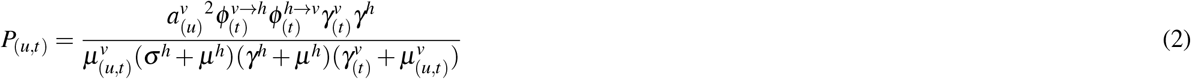

### Proposal of informative distributions of the biological parameters

The estimation of Index P requires some prior knowledge of the biology of the selected host, vector and viral system. In this study, we defined probability distributions for human hosts, *Ae. aegypti* vectors and dengue virus. Several mosquito species in the genus *Aedes* are known to transmit DENV to humans. We focused on *Ae. aegypti* because it accounts for the majority of vector-human transmission^39^, and it is the species for which the most empirical data exists on the relationship between climate and vector-viral traits. After a review of relevant literature, we defined probability distributions for six host-virus and vector-virus parameters: extrinsic *Ae. aegypti*-DENV incubation period, adult *Ae. aegypti* lifespan, adult *Ae. aegypti* biting rate, human lifespan, intrinsic human-DENV incubation period, and human-DENV infectious period (Table 1). These probability distributions are either directly sampled (*µ*^*h*^, *γ*^*h*^, *σ*^*h*^ and 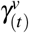) or used as likelihood functions for the estimation of climate-dependent probability distributions (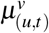 and 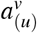).

### Climate-dependent functions for entomological parameters

Weather-dependent functions are defined for each of the entomological parameters in the expression for Index P (equation (2)). The adult human mortality rate (*µ*^*h*^), intrinsic incubation period (1/*γ*^*h*^) and the human infectious period (1/*σ*^*h*^) are taken to be climate-independent.

As detailed in Obolski et al.^24^, adult vector mortality (*µ*^*v*^) and the probability of transmission from vector to human per bite (*ϕ*^*v*→*h*^) are modeled by temperature-dependent functions estimated in experiments on laboratory strains of *Ae. aegypti* by Yang et al.^40^ (equation (3)) and Lambrechts et al.^41^ (equation (4)), respectively. In the original implementation of Index P, the probability of transmission from human to mosquito per bite (*ϕ*^*h*→*v*^) was assumed to be constant. To account for the temperature dependence of *ϕ*^*h*→*v*^ and to capture threshold effects at extreme temperatures, we expanded the framework by modeling *ϕ*^*h*→*v*^ with a ramp model estimated by Lambrechts et al.^41^ (equation (5)).

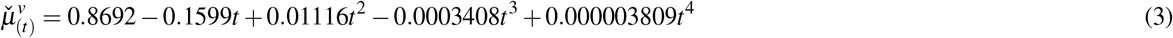

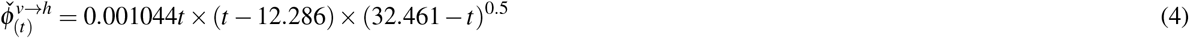

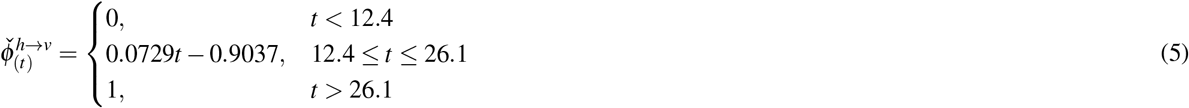

Following the original approach by Obolski et al.^24^, the influence of humidity on adult mosquito mortality rate (*µ*^*v*^) and mosquito biting rate (*a*^*v*^) is introduced according to the following expressions. The time series for relative humidity is normalized to [0, 1] with the ecological variables centered around the local average (*ū*).

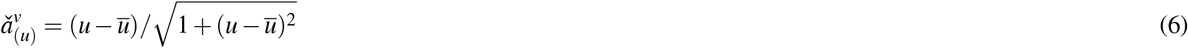

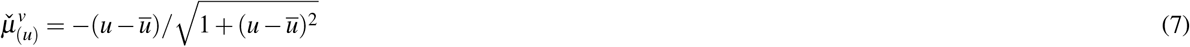

The total effect of climate on the entomological parameters is then modeled by combining the estimated temperature-dependent and humidity-dependent functions.

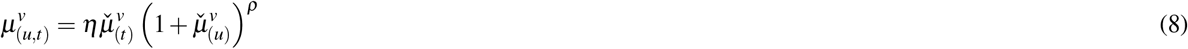

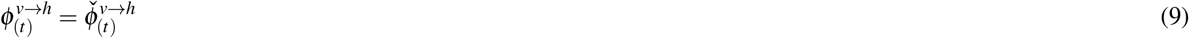

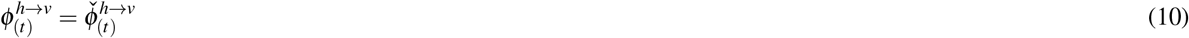

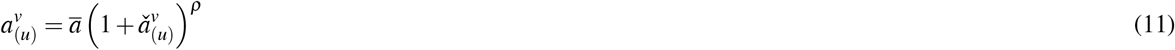

The scaling coefficients *η* and *ρ* in the expressions for mosquito biting rate (equation (11)) and adult mosquito mortality rate (equation (8)) are estimated from the user-defined probability distributions of *a*^*v*^ and *µ*^*v*^, respectively. These distributions permit deviations from the ideal laboratory conditions used in the original empirical studies. The multiplicative coefficient *η* determines the magnitude by which the effect of temperature on the adult mosquito mortality deviates from that observed under laboratory conditions. The effect of temperature under field conditions is identical to that under laboratory conditions when *η* = 0, stronger when *η >* 1 and weaker when *η <* 1. The exponential coefficient *ρ* modulates the relative influence that humidity has on adult mosquito mortality and mosquito biting rate: humidity has no effect when *ρ* = 0 and a stronger effect when *ρ >* 0. For further information on the biological interpretation of these coefficients and their estimation, please refer to Obolski et al.^24^.

Using recent, high-resolution empirical data, we also updated the probability distribution of the extrinsic incubation period (EIP) of *Ae. aegypti* mosquitoes. We introduced a temperature-dependent probability distribution of *γ*^*v*^ derived from observational studies of EIP in human subjects experimentally infected by wild *Ae. aegypti* mosquitoes^20^. Chan and Johansson^20^ estimate the temperature-dependent distribution of the EIP of *Ae. aegypti* mosquitoes (1*/γ*^*v*^) by directly fitting a log-normal time-to-event model to EIP observational data:

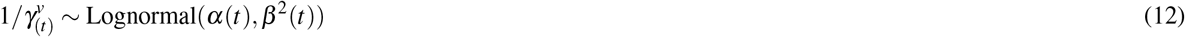

Where *α*(*t*) and *β* (*t*) are temperature-dependent functions of the mean and standard derivation of the natural logarithm of the EIP (Table 1).

### Estimation of Index P

Index P estimation is based on a Markov chain Monte Carlo (MCMC) sampling framework which derives the distribution of the Index P time series using the climate-driven functions for the entomological parameters (*µ*^*v*^, *ϕ*^*v*→*h*^, *ϕ*^*h*→*v*^, *a*^*v*^), the user-defined biological probability distributions (Table 1), and the climate time series data. Broadly, the estimation of Index P can be broken down into three steps. First, the posterior distributions of *µ*^*v*^ and *a*^*v*^ are estimated by fitting the scaling coefficients *η* and *ρ* to the corresponding probability distributions proposed by the user and the time series of the climatic variables. For each iteration of the MCMC procedure, the scaling coefficients are sampled and the climate-driven time series of *µ*^*v*^ and *a*^*v*^ are estimated. The posterior probability of the resultant time series is assessed through the product of the user-defined likelihood functions of *µ*^*v*^ and *a*^*v*^ with the prior probabilities of those parameters. Then, for each time point in the climate time series, the user-defined distributions of *µ*^*h*^, *γ*^*h*^, *σ*^*h*^ and *γ*^*v*^ and the derived posterior distributions of *µ*^*v*^ and *a*^*v*^ are independently sampled, and the temperature-dependent equations for *ϕ*^*v*→*h*^ and *ϕ*^*h*→*v*^ are solved. Finally, the sampled parameter values are plugged into the Index P expression, which yields a distribution of Index P time series bound according to the user-defined biological characteristics of the virus, vector, and host under investigation. An example of the workflow for the estimation of Index P time series at a given spatial pixel is provided in Figure 1.

**Figure 1.**
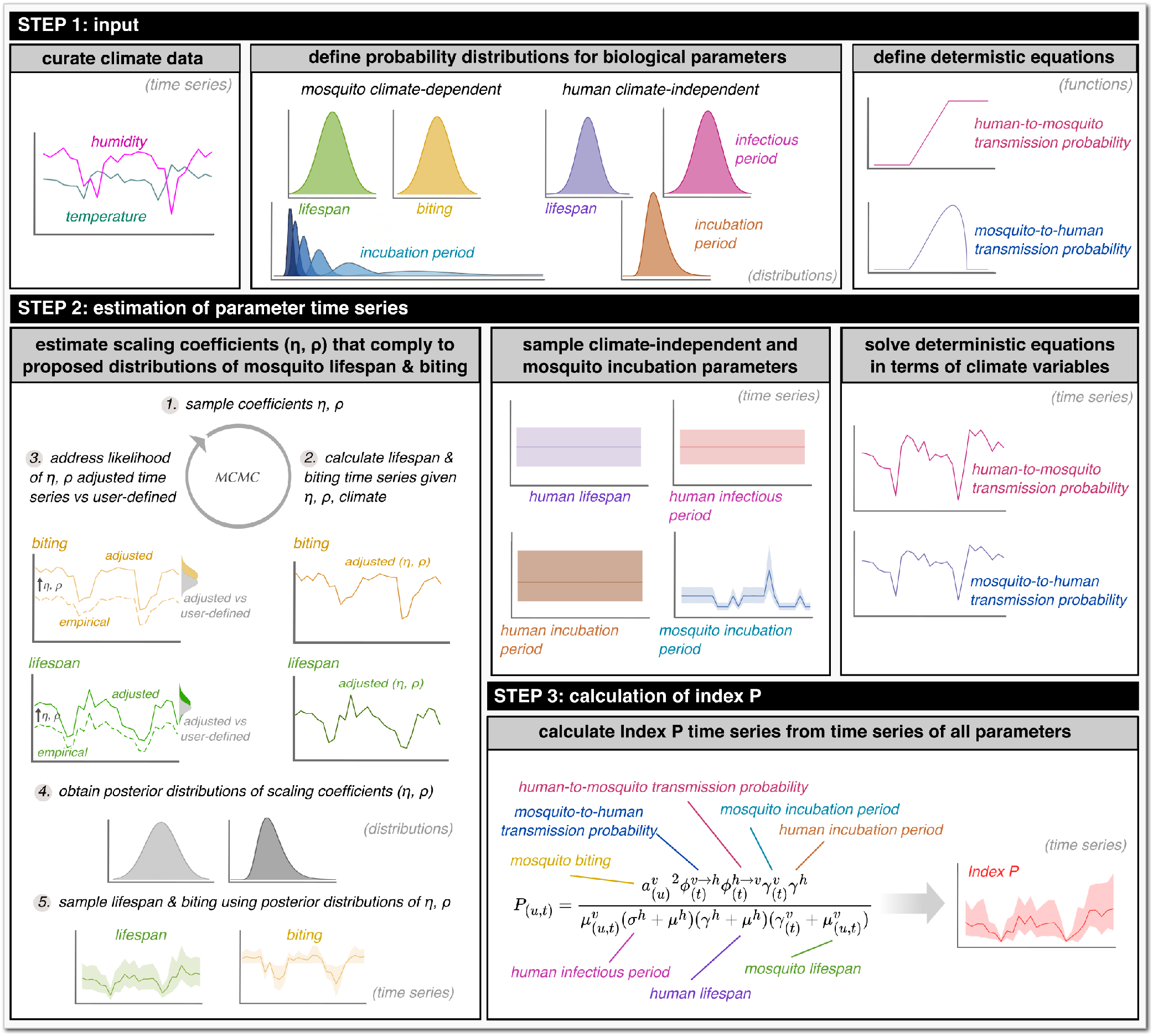
Summary of the steps required to estimate Index P at a single spatial pixel.

### Estimation and sampling of Index P globally

We used monthly temperature and humidity data, as well as informed probability distributions of the biological parameters as inputs to estimate the monthly Index P time series for each spatial pixel (∼ 28 km^2^) of 186 individual countries and territories from 1981 to 2019. For each country, spatial polygons were used to curate the monthly temperature and relative humidity data sourced from Copernicus.eu^42^ for all spatial pixels within its geographical boundary. The following steps were then performed for each spatial pixel. Using the climate data and the user-defined probability distributions for mosquito biting rate (*a*^*v*^) and adult mosquito mortality rate (*µ*^*v*^), we estimated the climate-dependent posterior distributions of these two parameters by fitting the scaling coefficients *η* and *ρ*. For each MCMC chain, we drew 20, 000 samples with the first 20% of samples treated as burn-in. Given a lack of information on the possible estimates of *ρ* and *η*, we assumed uninformative, flat priors for the two scaling coefficients in the ranges 0 − 10 and 0 − 20 respectively. We then drew 1, 000 samples from the climate-independent distributions of *µ*^*h*^, *γ*^*h*^ and *σ*^*h*^, 1, 000 samples from the climate-dependent distributions of the *γ*^*v*^, *µ*^*v*^ and *a*^*v*^ time series, and solved the deterministic temperature-dependent equations of *ϕ*^*v*→*h*^ and *ϕ*^*h*→*v*^. These sampled values were plugged into the equation for Index P, resulting in 1, 000 monthly Index P time series. Finally, we calculated various summary statistics of the distribution of monthly Index P time series for each spatial pixel. Details on the calculated summary statistics are provided in the Data Records section. These estimates were collated at the country/territory level into TIFF files^43^.

## Data Records

### Meteorological data

The meteorological data from 1981 to 2019 that were used to estimate the spatiotemporal maps of Index P are freely available from the “Essential Climate Variables for assessment of climate variability from 1979 to present” dataset published and maintained by Copernicus.eu^42^. We used the average monthly surface air temperature (*K*) and surface air relative humidity (%). The spatial pixel resolution used was 0.25° ×0.25° (∼28 km^2^), the maximum resolution available for the Copernicus.eu dataset.

### DENV case data

Weekly notified dengue case data per municipality in Brazil for the years 2000 to 2014 were provided by the Brazilian Ministry of Health (https://www.gov.br/pt-br) and by the SINAN (Sistema de Informação de Agravos de Notificação; http://portalsinan.saude.gov.br/)^44^. It includes clinically suspected infections without laboratory confirmation for 5,570 municipalities. Four municipalities had incomplete time series data and were excluded from the analysis: Mojuí dos Campos (Pará), Pescaria Brava (Santa Catarina), Balneário Rincão (Santa Catarina) and Paraíso das Águas (Mato Grosso do Sul).

### Population data

The yearly population size per municipality in Brazil for the years 2001 to 2014 was sourced from IBGE - Instituto Brasileiro de Geografia e Estatística (https://www.ibge.gov.br/en/home-eng.html)^45^. Missing population data for the year 2000 was assumed to be the same as that for the year 2001.

### Global Index P data

For each country or territory, we calculated epidemiologically relevant summary statistics for the spatiotemporal distribution of Index P. These include: spatiotemporal map(s) for each month (468 layers), each year (39 layers), a typical year (12 layers) and the entire period (1 layer); spatiotemporal maps of the number of months Index P is above 1.0 for each year (39 layers), and for a typical year (12 layers); and spatiotemporal maps of the peak and trough timing during a typical year (12 layers each). The spatiotemporal maps are provided as TIFF files^43^. The Index P spatiotemporal maps are projected with World Geodetic System (WGS)84, latitude/longitude coordinate system (EPSG: 4326; https://epsg.io/4326). The complete dataset and a description of its contents are available from a figshare repository (http://www.figshare.com/)^43^. The Index P maps can be processed with packages such as *raster*^46^ in the R programming language. A sample of the Index P maps and time series is in Figure 2.

**Figure 2.**
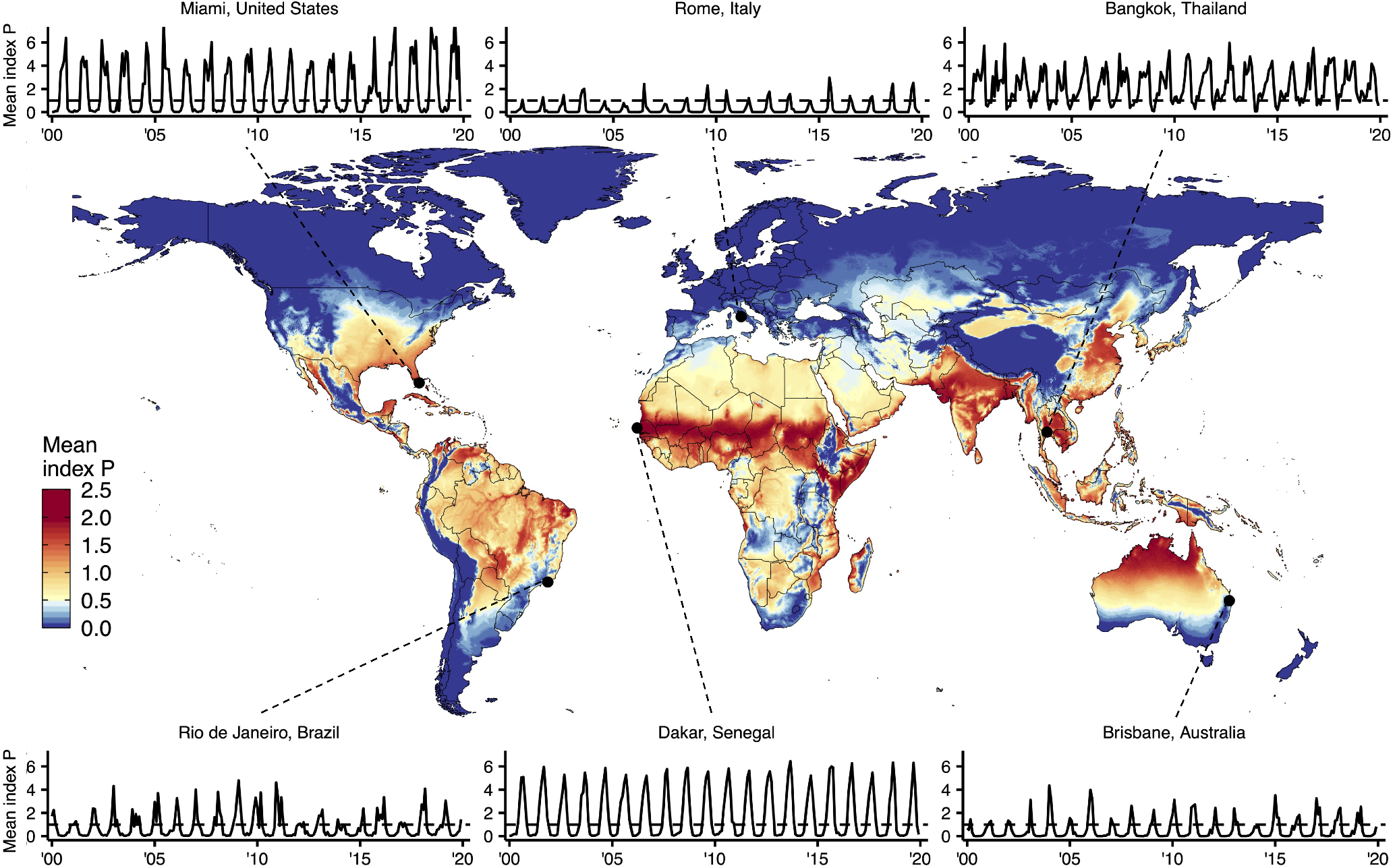
Summary of global spatiotemporal maps of estimated Index P for DENV transmitted by *Aedes aegypti* mosquitoes. Global map of mean Index P during a typical year at a spatial resolution of 0.25° ×0.25° (∼28km^2^). Includes monthly time series of average monthly Index P from 2000 to 2019 for six cities: Bangkok, Thailand; Brisbane, Australia; Dakar, Senegal; Miami, United States; Rio de Janeiro, Brazil; and Rome, Italy. Cities were selected to capture the diversity of Index P dynamics observed globally across regions where DENV is endemic, emerging or absent.

### Technical Validation

Using historical DENV case data recorded in Brazil from 2000 to 2014, we performed a validation of Index P as a climate-based measure of transmission potential. We focused on Brazil for several reasons: (1) DENV is endemic with high disease burden^8, 47^, (2) the country is climatically diverse with both temperate and tropical areas^48^ and there is large variation in the intensity and seasonality of observed DENV dynamics^49^. Given that DENV incidence data are available at the municipality level for Brazil, we calculated and validated the relevant summary statistics of Index P at the municipality level rather than at the spatial pixel level. Here, we included several analyses that support the application of Index P to explain past spatiotemporal dynamics of DENV, reaffirming the results of previous studies^5, 24, 26–32^.

### Local transmission intensity

To quantify how well the values of Index P characterize spatial variation in local DENV incidence, we compared the mean Index P during a typical year and the mean yearly incidence in each municipality across Brazil. A typical yearly time series is estimated by calculating the mean value of a chosen variable (e.g. Index P, log(incidence+1), incidence, etc.) independently for each of the twelve months of the year across all available years. The map of transmission potential was consistent with the spatial distribution of incidence (Figure 3a-b). The hyperendemic regions in the Northeast and the Midwest had higher estimated transmission potential, while the low incidence areas in the South stretching from Rio Grande do Sul to Minas Gerais had lower estimated transmission potential.

**Figure 3.**
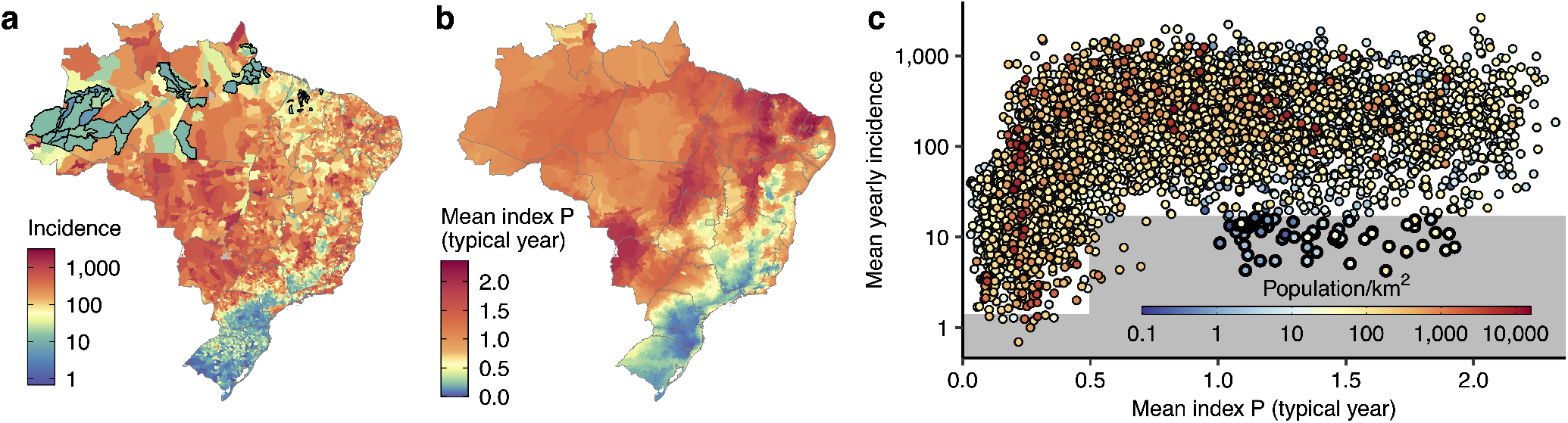
Comparison of reported DENV incidence and mean Index P for a typical year (**a**) Mean yearly incidence in each municipality. (**b**) Mean annual Index P for a typical year. (**c**) The relationship between mean annual Index P and mean yearly incidence. The area below the lower fence (*Q*1 − 1.5 * *IQR* for (0, 0.5) and (0.5, ∞)) which encompasses outliers is shaded grey with municipalities that form part of Brazil’s Legal Amazon highlighted black (these municipalities are also highlighted in panel **a**).

Estimated transmission potential and incidence exhibited a nonlinear relationship. Incidence increased with Index P until reaching a plateau in which increases in transmission potential did not yield higher incidence (Figure 3c). This nonlinear relationship is consistent with the interpretation of Index P as the absolute transmission potential of an adult mosquito, where increases in Index P are tied to increases in *R*_0_ and the probability and scale of epidemic growth^50^. The observed plateau at high Index P values can be explained by the dynamics of viral transmission, where the size of epidemics is constrained by factors other than transmission potential, particularly the development of herd immunity over time. As such, municipalities estimated to have very high transmission potential presented similar levels of high incidence.

The reported incidence was lower than expected from the estimated transmission potential in several municipalities in the Northwest states of Amazonas and Pará (Figure 3a-b). Given that these areas form part of the Amazon rainforest, these inconsistencies might be due to a combination of poor surveillance, lack of connectivity to allow frequent viral introduction and/or low population density. Indeed, the municipalities within Brazil’s Legal Amazon that had lower than expected incidence had some of the largest areas of all municipalities, and relatively low population densities (Figure 3A, 3c).

### Typical year seasonality

We compared the Index P dynamics to the observed DENV seasonality by calculating Spearman’s correlation coefficient between the time series of Index P and log-transformed incidence in each municipality during a typical year (Figure 4). Municipalities with an average of fewer than 12 cases per year (2725/5570) were excluded since their incidence dynamics did not have sufficient information for a measurable seasonal signal. To quantify the time delay between the Index P and incidence dynamics, we calculated the correlation coefficient for all possible monthly lags (i.e. -5 to 6 months inclusive) and selected the lag with the highest coefficient. The lag with the highest correlation coefficient and the coefficient itself are referred to as the lag and lag-adjusted correlation of a municipality respectively.

**Figure 4.**
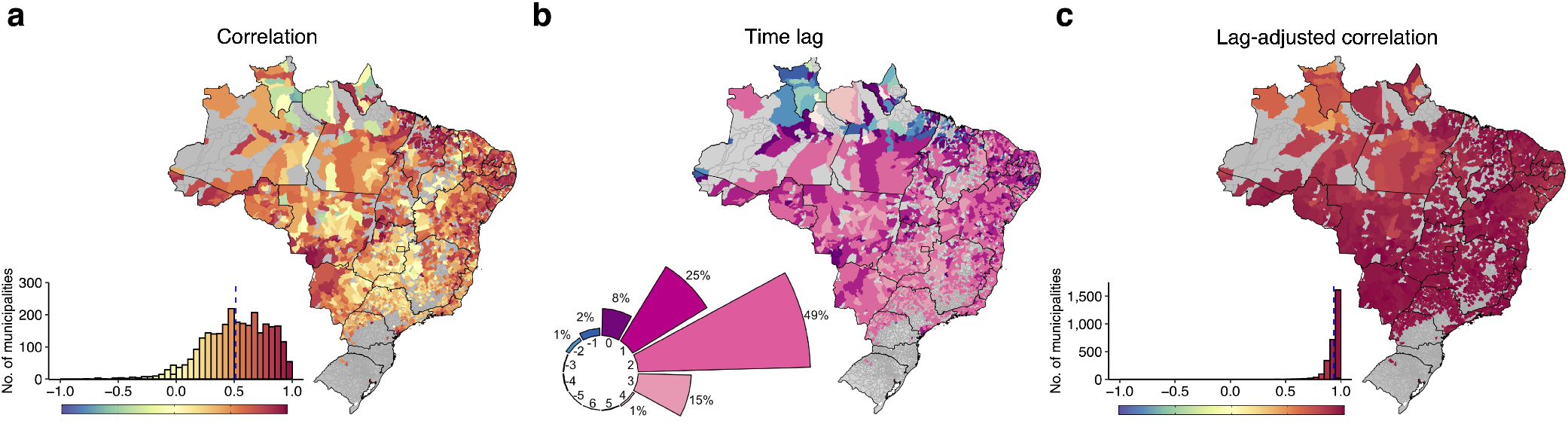
Correlation between the time series of Index P and incidence during a typical year at the municipality level. (**a**) Lag-unadjusted Spearman’s correlation coefficient between Index P and log-transformed incidence during a typical year. The histograms show the distribution of coefficients (mean represented by blue dashed line). Municipalities with fewer than 12 cases per year on average are excluded and coloured grey in the maps (N=2725) given that they have insufficient case data to have detectable seasonal signals. (**b**) Map of the predicted lag of each municipality. The circular barplot shows the distribution of lags from -5 months to +6 months. The proportion of municipalities observed for each lag is noted (proportions less than 1% are not shown). (**c**) Lag-adjusted Spearman’s correlation coefficient between Index P and log-transformed incidence during a typical year.

The distribution of lag-unadjusted correlation coefficients was left skewed with 42% of municipalities showing relatively high coefficients greater than 0.6 (Figure 4a). Given the development time of *Aedes* mosquitoes (∼2 weeks), the generation time of DENV in human populations (∼2 −3 weeks)^51, 52^and the time for mosquitoes to respond to climatic signals, it can take many weeks for increases in transmission potential to translate into increases in incidence^5, 53, 54^. Indeed, the Index P dynamics in 88% of municipalities preceded the incidence dynamics by 1-3 months, with the proportion of municipalities with time delays of 1, 2 and 3 months being 25%, 47%, and 15% respectively (Figure 4b). When we corrected for this time delay, the dynamics for Index P and incidence were highly correlated with 82% of the municipalities with an average of 12 or more cases per year showing correlation coefficients greater than 0.90 (Figure 4c).

Some municipalities (4%) had extreme lags (i.e. negative or greater than 3 months) inconsistent with the biological interpretation of Index P (in the North, spanning the states of Roraima, Pará, Amapá and Maranhão, Figure 4b). Given that these regions have monsoon-driven climates characterized by intense timely annual rainfall, a factor not considered in Index P but known to influence mosquito abundance, precipitation may play an outside role in determining seasonality in these regions.

### Usage Notes

Researchers can use Index P maps to explore monthly transmission potential of DENV spread by *Ae. aegypti* mosquitoes at the spatial pixel level (∼28 km^2^) from 1981 to 2019. There are, however, several caveats to the application of these maps. Firstly, from our technical validation, it can be concluded that areas with low estimated transmission potential can be expected to have lower incidence. However, areas with high estimated potential are susceptible to more uncertainty, as non-climatic factors, particularly accumulated herd immunity and host/vector densities, can influence whether favorable climatic conditions translate into higher disease incidence. High Index P thus should not be interpreted as a guarantee of transmission but rather as one indicator of favorable climatic conditions for DENV circulation. Secondly, while Index P dynamics reliably capture the seasonality of dengue transmission, there is likely to be a time delay between the proposed climatic effects and the observed changes in incidence, the length of which can vary between locations. Our analyses, and those of others^53, 54^, suggest that this time delay is between about 1 to 3 months, but a comparison of past incidence and Index P dynamics at a local level is necessary for more precise, locally-specific estimates. Finally, Index P is unlikely to mimic incidence patterns at very high spatiotemporal resolutions due to obfuscation of the seasonal signals of incidence by factors such as imperfect surveillance and local stochasticity. Thus Index P is most informative at aggregated spatiotemporal resolutions where incidence is expected to demonstrate clear seasonal signals.

To encourage reuse and deepen understanding of the Index P methodology, we developed a new version of the **M**osquito-borne **V**iral **S**uitability **E**stimator (MVSE) software package for the R programming environment^24^. This R package provides a set of related functions that can be used to estimate Index P time series given temperature and humidity time series and user-defined probability distributions of the biological parameters for the selected host/vector/virus system. A short tutorial on the features of the MVSE package is also made available. Furthermore, we have provided an R Markdown document that highlights useful R software packages and functions for the visualization and analysis of the dataset of Index P maps. Though all code used to generate this dataset was written in R and C++, the maps themselves are provided as TIFF files to facilitate cross-compatibility.

## Data Availability

The meteorological data from 1981 to 2019 that were used to estimate the spatiotemporal maps of Index P are freely available from the Essential Climate Variables for assessment of climate variability from 1979 to present dataset published and maintained by Copernicus.eu (https://cds.climate.copernicus.eu/cdsapp#!/dataset/ecv-for-climate-change).
Weekly notified dengue case data per municipality in Brazil for the years 2000 to 2014 were provided by the Brazilian Ministry of Health (https://www.gov.br/pt-br) and by the SINAN (Sistema de InformaÇão de Agravos de NotificaÇão -  http://portalsinan.saude.gov.br/).
The yearly population size per municipality in Brazil for the years 2001 to 2014 was sourced from IBGE - Instituto Brasileiro de Geografia e Estatística (https://www.ibge.gov.br/en/home-eng.html).
The complete dataset of spatiotemporal index P maps and a description of its contents are available from a figshare repository at https://doi.org/10.6084/m9.figshare.21502614.

https://www.ibge.gov.br/en/statistics/downloads-statistics.html

http://portalsinan.saude.gov.br/

https://doi.org/10.6084/m9.figshare.21502614

## Code availability

The MVSE R package can be installed from the GitHub repository at https://github.com/TaishiNakase/MVSE. The MVSE package tutorial and the R Markdown document that provides some example code for the visualization of the Index P maps can be found at the GitHub repository https://github.com/TaishiNakase/Index-P-estimation-and-applications.

## Author contributions statement

T.N. generated and analyzed the data, wrote the code and led the writing of the manuscript. J.L. conceived the idea, supervised the project and contributed extensively to all ideas and writing. M.G. and U.O. read and critically revised the manuscript.

